# Modeling the Coronavirus Disease 2019 Incubation Period: Impact on Quarantine Policy

**DOI:** 10.1101/2020.06.27.20141002

**Authors:** Daewoo Pak, Klaus Langohr, Jing Ning, Jordi Cortés Martínez, Guadalupe Gómez Melis, Yu Shen

## Abstract

The incubation period of coronavirus disease 2019 (COVID-19) is not always observed exactly due to uncertain onset times of infection and disease symptom. In this paper, we demonstrate how to estimate the distribution of incubation and its association with patient demographic factors when the exact dates of infection and symptoms onset may not be observed. The findings from analysis on the confirmed COVID-19 cases indicate that age could be associated with the incubation period, and an age-specific quarantine policy might be more efficient than a unified one in confining COVID-19.

## 1. Introduction

The coronavirus disease 2019 (COVID-19) was first reported in Wuhan, China, in December 2019; an outbreak rapidly spread worldwide. The novel virus infection can be asymptomatic or unapparent during a certain period and asymptomatic persons could spread the virus unknowingly [1]. Among patients who develop symptoms, the incubation period is defined as the elapsed time between infection and appearance of the first symptom. Knowledge of the incubation period is essential for disease prevention, facilitating an optimal quarantine guideline to confine the spread.

The distribution of incubation for coronavirus disease 2019 (COVID-19) has been investigated in several reports. In the early outbreak, Backer et al. [2] estimated the incubation period distribution among travelers from Wuhan, and Linton et al. [3] investigated the geographic spread of the infections from Wuhan. Lauer et al. [4] estimated the distribution among confirmed cases outside Wuhan. The demographic and clinical characteristics of COVID-19 in China were also discussed in Guan et al. [5]. Despite its importance, it remains unclear how the incubation distribution could vary by age and gender. The current 14-day quarantine period, ignoring the patient demographic factors, may be insufficient for the containment of COVID-19. Only a few studies [6, 7] addressed these concerns using a limited amount of confirmed cases. A more accurate estimation of the incubation period using these factors is necessary to optimize the surveillance guidelines.

We model uncertain dates of infection and symptom onset with the best available information, by classifying COVID-19 confirmed cases into plausible observing scenarios. The observed data are subject to both right-censoring and left-censoring; if these censoring mechanisms are not properly considered, the estimates for the incubation period could lead to severe biases. Ignoring the left-censoring tends to overestimate the incubation period. This approach allows the use of incomplete observations on the COVID-19 incubation period and thereby provides more reliable results and inferences. A sufficiently general parametric class, the generalized odds-rate class of regression models, is employed for modeling the incubation period of COVID-19. This parametric class includes the log-logistic proportional odds model and the Weibull proportional hazards model as special cases. This modeling is more flexible than the distributions commonly used in the previous relevant work [2, 3, 4, 5, 6, 7, 8]. We also revisit the impact of the current quarantine duration on the spread of COVID-19 by estimating the distribution of the incubation periods of COVID-19 and its association with gender and age.

## 2. Exposure History Reports for Confirmed COVID-19 Cases

We outline a modeling approach to enable the analysis of publicly reported, clinically confirmed cases with symptoms from two sources [9, 10], available as of March 30, 2020. The publicly available data sets for these two sources are freely available without permission requirement by DXY [9] (https://docs.google.com/spreadsheets/d/e/2PACX-1vQU0SIALScXx8VXDX7yKNKWWPKE1YjFlWc6VTEVSN45CklWWf-uWmprQIyLtoPDA18tX9cFDr-aQ9S6/pubhtml) and Xu et al. [10] (https://github.com/beoutbreakprepared/nCoV2019/tree/master/covid19/raw-data), respectively. To obtain the period of potential exposure, defined as contact with infected persons or visit to contaminated regions, we included the following patients in the analytic data set: 1) non-residents of Wuhan who visited Wuhan since December 1, 2019 and for whom the exposure period was the time between the earliest possible arrival to and the latest possible departure from Wuhan; 2) people with a travel history of visiting the coronavirus-affected nations known at the time or taking the Diamond Princess Cruises; 3) non-travel related cases with an exposure history based on contact with infected persons. Among these, 312 cases had recorded exposure periods, or at least an exposure end date, and dates of symptom onset or hospitalization, as well as gender and age; this number reflects removal of 34 potentially duplicated cases (see Figure 1).

**Figure 1:**
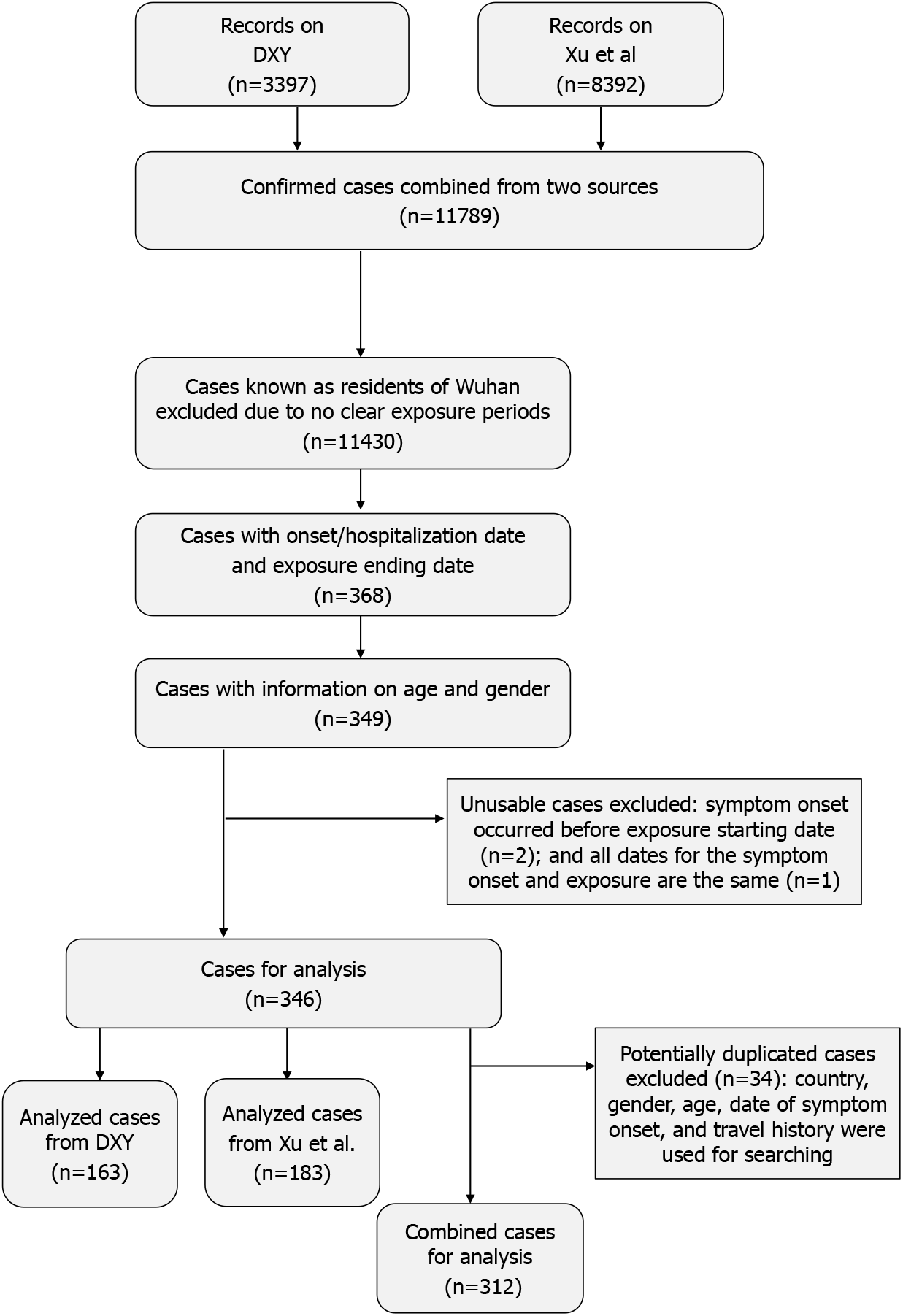
Flow diagram of confirmed cases for data analysis. Two data sources were combined from DXY [9] and Xu et al [10].

There were 111 (35.6%) cases with known exposure start and end dates, illustrated in Supplementary Figure S1. For cases without an exact start date of the exposure period, the initial date was set to have a maximum of thirty days of exposure or was set to December 1, 2019, whichever one was later. In addition, the end date of exposure was set to precede the known dates of symptom onset and hospitalization. If the exact date of symptom onset was unknown, it was assumed to have occurred before hospitalization (i.e., left-censored).

## 3. Model and Estimation

The distribution of the COVID-19 incubation period is estimated with the patient-related covariates by modelling the interval-censored exposure duration and the possibly left-censored symptom onset time. The proposed method allows us to use a larger data cohort consisting of more confirmed COVID-19 cases; the cases may not have the exact symptom onset time (left-censored) and include an interval-censored incubation period. We parameterize the underlying incubation period under the generalized odds-rate class of regression models with the patient’s age and gender. Let *T* be the incubation period and ***X*** be the covariate vector including both the patient’s age and gender. Within the generalized odds-rate class of regression models, the conditional probability density function of *T* given ***X*** = *x* is defined as

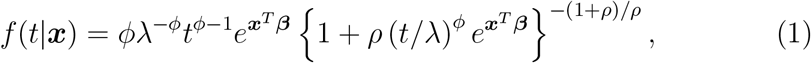

where ρ, λ, and *ϕ* are the positive model parameters, and *β* is the vector of the regression parameters. The corresponding survival function is

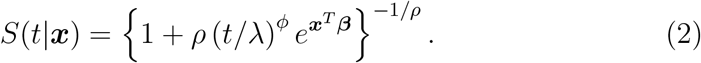

This class of models includes the log-logistic proportional odds model (*ρ* = 1) and the Weibull proportional hazards model (*ρ* → 0) as special cases [11], where survival functions are respectively expressed as

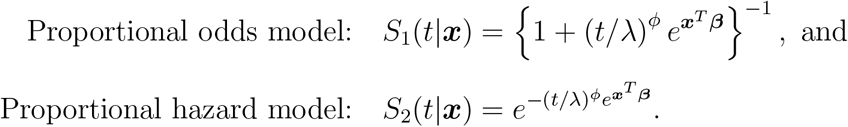

To construct the likelihood function of COVID-19 data from (1) and (2), we distinguish the four types of observation, as shown in Figure 2. Let d denote the difference between the exposure start date and end date, and r denote the difference between the exposure end date and either the symptom onset date (if available) or the date of hospitalization. If the date of infection is at best known to be within a potential exposure period (i.e., *d* > 0), let *δ_d_* =1; otherwise *δ_d_* = 0. If a patient’s symptom onset date was observed, let *δ_s_* = 1; otherwise, *δ_s_* = 0. Under the assumption of a uniform risk for infection during the potential exposure periods, the contributions of each type to the likelihood function, defined as *L*_1_, *L*_2_, *L*_3_, and *L*_4_, respectively, are given as follows:

- Type I: observing potential exposure period and symptom onset date (*δ_d_* = 1 and *δ_s_* = 1)

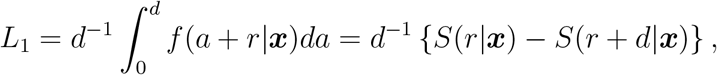
- Type II: observing potential exposure period and hospitalization date (*δ_d_* = 1 and *δ_s_* = 0)

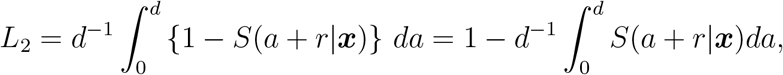
- Type III: observing exact exposure date and symptom onset date (*δ_d_* = 0 and *δ_s_* = 1)

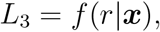
- Type IV: observing exact exposure date and hospitalization date only (*δ_d_* = 0 and *δ_s_* = 0)

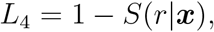

where *a* is the variable of integration representing the unknown period from the infection to the exposure end.

**Figure 2:**
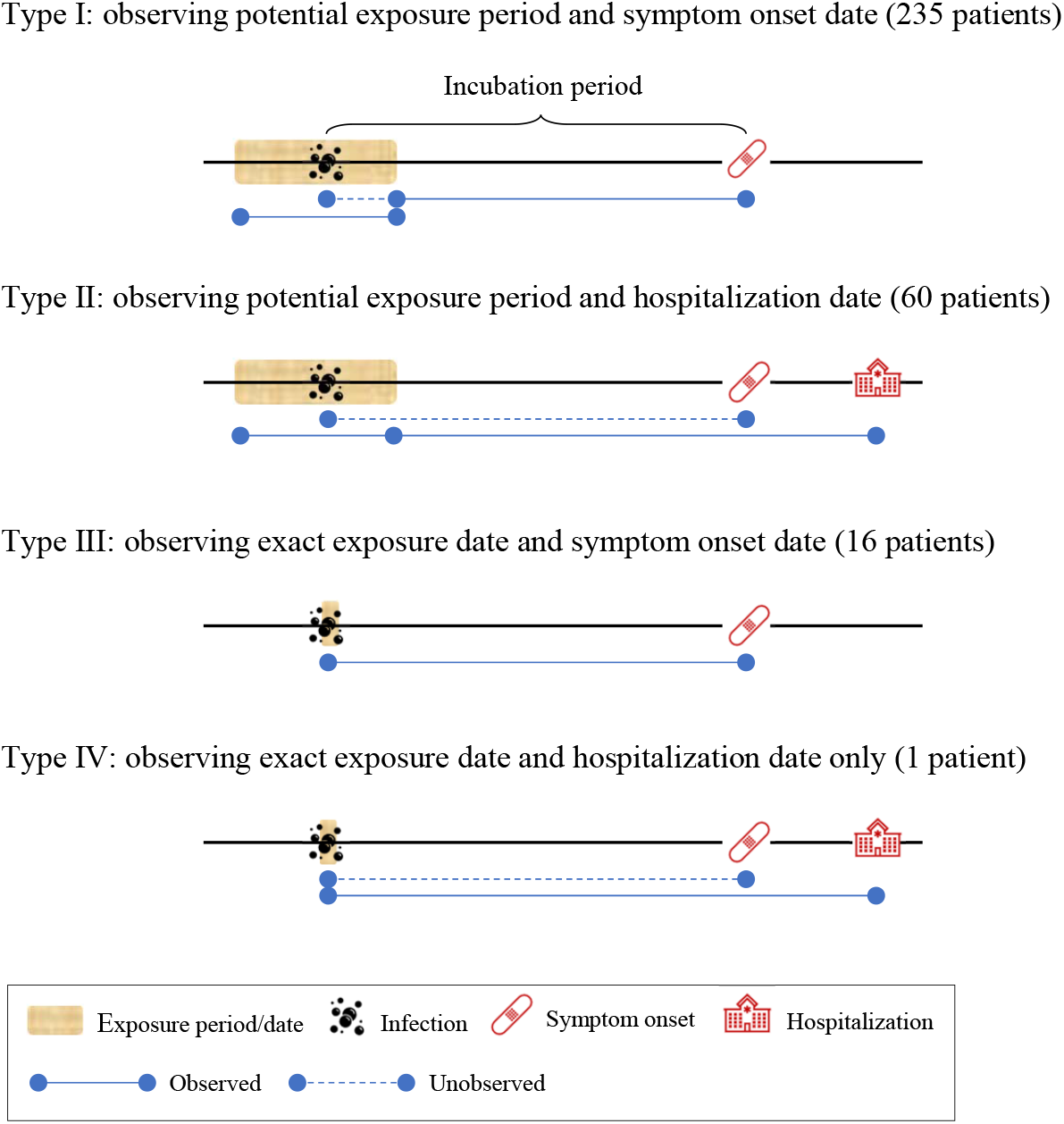
The four types of observations available for the data analysis. The total number of patients is 312.

Considering *n* independent observations, the overall log-likelihood is then proportional to

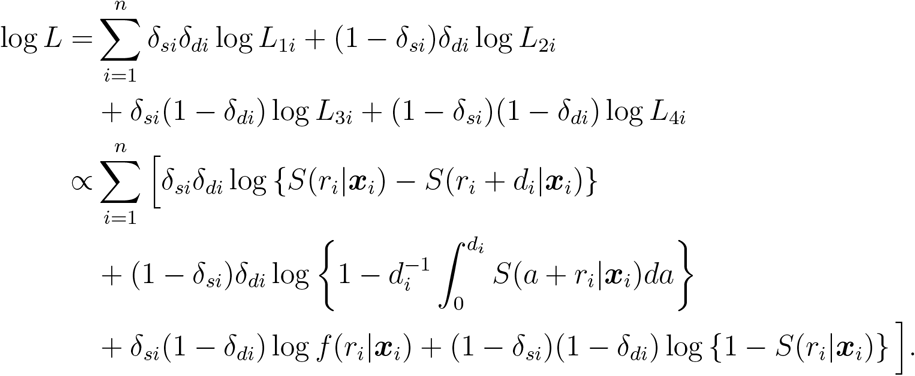

The model parameters are estimated by maximizing the log-likelihood. In our implementation, we maximized the log-likelihood with the logarithmic transformation for the positive parameters (i.e., *λ*, *ϕ*, and *ρ*). The delta method was used to obtain the asymptotic normal distribution for any parameter transformation, such as the median incubation time. All analyses were performed with R, version 3.6.1 [12]. The R code is provided with the supplementary material.

## 4. Application

We applied the proposed method to the COVID-19 data sets, described in Section 2. Within the total of 312 patients, the median age was 42 (interquartile range 33 – 55) years, and 126 (40.4%) patients were women. The summary statistics and the histogram for patient’s age are shown in Supplementary Table S1 and Figure S2. The positive association between age and the duration of incubation was observed from the model with age as a continuous variable (Supplementary Table S2). Age was dichotomized at its median prior to the analysis. Table 1 shows the estimation results for regression models with gender and age. An interaction between age and gender was not statistically significant in the model. Patients older than 42 years of age have, on average, longer incubation periods, compared to 42-year-old or younger patients (*p* = 0.039), whereas gender has no effect on the incubation period (*p* = 0.423). We decide not to include gender in the final model (Reduced model of Table 1), based on the likelihood ratio test with *p* = 0.384. For the outbreak period from December 2019 to March 2020, the median incubation times are estimated to be 5.3 (95% CI: 4.3 – 6.3) days and 7 (95% CI: 5.6 – 8.3) days among younger (≤ 42 years) and older patients (>42 years), respectively. During this outbreak period, the mean incubation period is estimated to be 6.6 (95% CI: 5.4 – 7.8) days for the younger patients, and 8.8 (95% CI: 7.2 – 10.3) days for the older patients. The estimated distribution functions in both age groups together with the 95% confidence intervals are presented in Figure 3. Table 2 shows the estimated differences (in days) between the quantiles of the incubation distribution in both age groups. The incubation period difference between the two age groups was not obvious in lower quantiles, but it becomes conspicuous after the 25th quantile, leading to about 7-day differences between the two age groups at the 97.5th quantile.

**Table 1:** Results of the analysis of COVID-19 under the generalized odds-rate class of regression models.

| Model | | $\log(\lambda)$ | $\log(\phi)$ | $\log(\rho)$ | $\beta_{male}$ | $\beta_{age \leq 42}$ |
| --- | --- | --- | --- | --- | --- | --- |
| Full model | Estimate | 2.300 | 0.305 | -3.766 | 0.141 | 0.371 |
|  | SE | 0.155 | 0.119 | 6.569 | 0.176 | 0.180 |
|  | P-value | <0.001 | 0.010 | 0.566 | 0.423 | 0.039 |
| Reduced model | Estimate | 2.115 | 0.457 | -1.271 | - | 0.449 |
|  | SE | 0.126 | 0.128 | 0.685 | - | 0.214 |
|  | P-value | <0.001 | <0.001 | 0.064 | - | 0.035 |

\* Full model refers to the model with age and gender and reduced model refers to the model with age only. The p-value of the likelihood ratio test comparing the full model and reduced model was 0.384.

**Figure 3:**
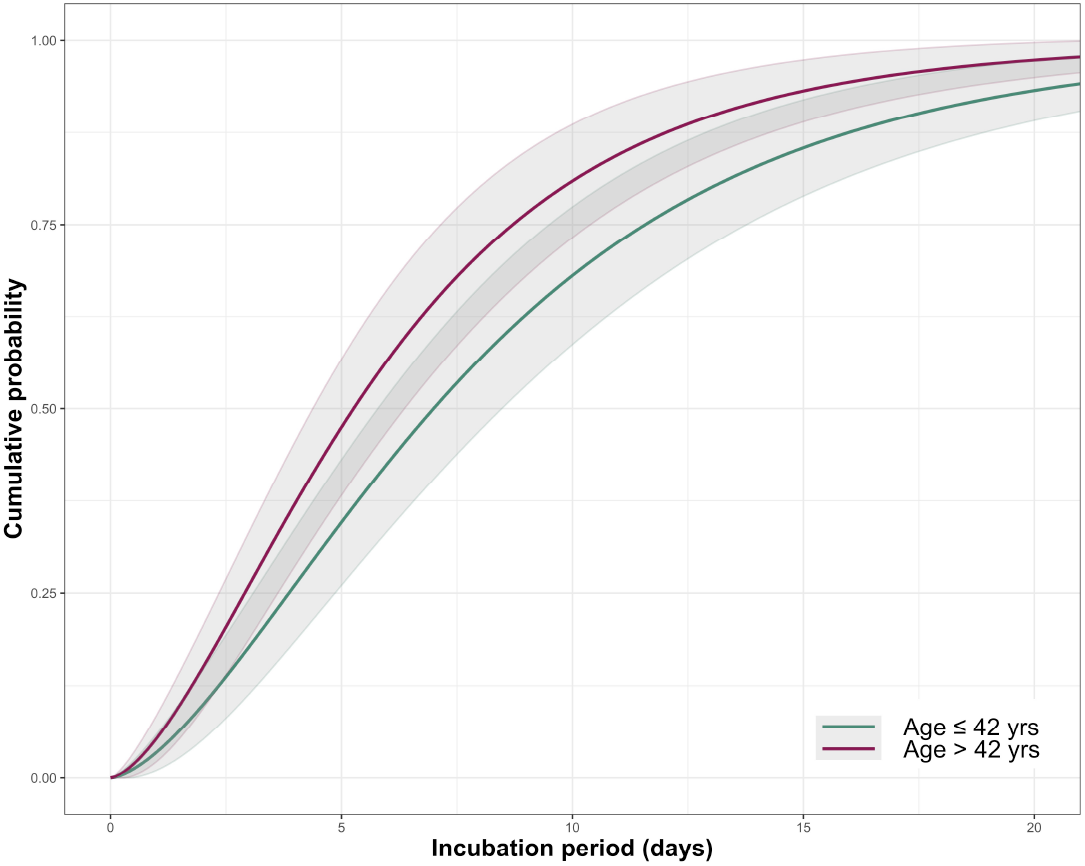
The estimated cumulative distribution of the incubation period by two age groups.

**Table 2:**
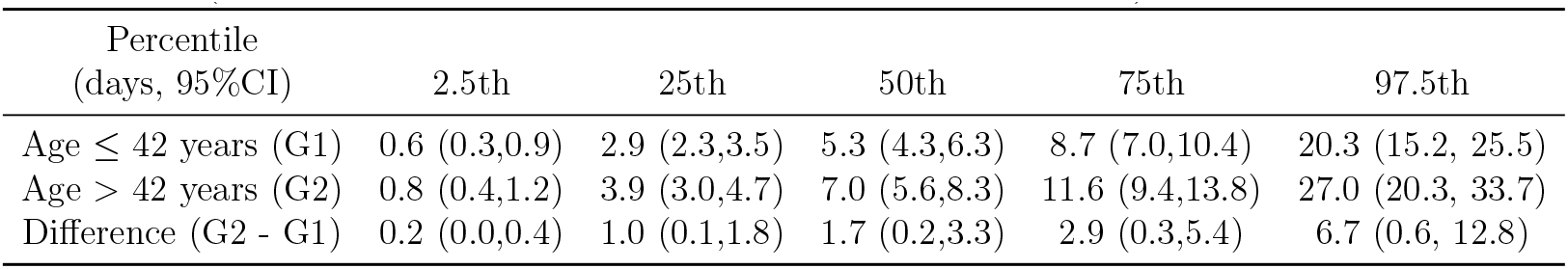
Estimated differences of specific quantiles of the incubation period among both age groups (patients aged > 42 years - patients aged ≤ 42 years).

| Percentile<br>(days, 95%CI) | 2.5th | 25th | 50th | 75th | 97.5th |
| --- | --- | --- | --- | --- | --- |
| Age $\leq 42$ years (G1) | 0.6 (0.3,0.9) | 2.9 (2.3,3.5) | 5.3 (4.3,6.3) | 8.7 (7.0,10.4) | 20.3 (15.2, 25.5) |
| Age $> 42$ years (G2) | 0.8 (0.4,1.2) | 3.9 (3.0,4.7) | 7.0 (5.6,8.3) | 11.6 (9.4,13.8) | 27.0 (20.3, 33.7) |
| Difference (G2 - G1) | 0.2 (0.0,0.4) | 1.0 (0.1,1.8) | 1.7 (0.2,3.3) | 2.9 (0.3,5.4) | 6.7 (0.6, 12.8) |

## 5. Discussion

The finding that persons older than 42 years have, on average, longer incubation periods than those who are younger may have important implications for enacting age-specific quarantine policies. This result agrees with previous studies on Severe Acute Respiratory Syndrome that showed a relationship between age and the incubation period [13]. Using the uniform 14-day quarantine policy recommended by the World Health Organization and the US Centers for Disease Control and Prevention [14, 15], our estimators imply that 8.4% (95% CI: 3.6 – 13.1%) of COVID-19 patients younger than 42, and 17.1% (95% CI: 9.9 – 24.2%) of older patients may pose a risk of infection to others before onset of their symptoms. Using a 21-day quarantine, these percentages reduce to 2.2% (95% CI: 0 – 7.0%) and 5.8% (95% CI: 1.3 – 13.0%), respectively. To ensure that at least 90% of cases’ symptoms are being manifested during quarantine periods, the required durations are estimated to be 13.1 (95% CI: 10.5 – 15.7) days for patients 42 years of age or younger and 17.4 (95% CI: 14.0 – 20.8) days for patients older than 42 years. As such, a unified quarantine policy could be inefficient during a viral outbreak. These estimates were derived from the conservative assumption that the quarantine periods started immediately after infection.

To examine how robust the estimates were to the assumption of 30 days of maximum exposure for cases with a missing start date of exposure interval, we performed some sensitivity analyses considering a maximum exposure duration of 20 days or shifting the lower bound back and forth within 15 days. The variation of these assumptions had little effect on analysis results (Supplementary Table S3). Although the proposed method assumes that the infection would happen uniformly during the potential exposure period, it can be easily modified with other forms of risk for various purposes.

The application to the COVID-19 data sets has some notable limitations. Our inferences relied on publicly reported confirmed cases that might over represent more severely symptomatic patients. Moreover, the definition of COVID-19 symptoms and hospitalization criteria could differ by country, especially during the initial outbreak. We combined the data sets from two different sources, and the potential variation in source criteria for tracing infected cases may lead to different exposure records. However, we obtained similar findings when fitting the model to each data set separately. The same trend was observed for the incubation period by the age groups (Supplementary Table S4 and Table S5), though one indicated no statistically significant difference. We dichotomized age to show the difference in the incubation period time that might exist between the two groups. However, in no case was it our goal to identify an optimal cut-off age, since we are aware of the risks involved in the dichotomization of the explanatory variables [17, 16].

The longer incubation periods experienced by older patients might have been due to a delayed immune response system, given the mechanism of immune systems against COVID-19 [18]. However, the results may not be directly applicable to affect the public health policy globally, because the distribution of the incubation period could differ by other factors such as case reporting system, and co-infection levels in different regions and countries. There is also a possibility that the virus evolves to be more or less lethal and transmissible over time [19]. As one referee commented, co-morbidities and medical history would be informative to further investigate their association with the incubation period of COVID-19. They could be directly incorporated in our model as covariates. Unfortunately, our data sources do not include such information; thus, we cannot investigate those effects on the incubation period.

## Data Availability

Our analysis was based on publicly reported, clinically confirmed cases with symptoms from two sources, available through the links in the bibliography.

## Author Contributions

Conceptualization, D.P., G.G., J.N. and Y.S.; Methodology, D.P., J.N. and Y.S.; formal analysis, D.P., K.L., J.N. and J.C.; writing-original draft preparation, D.P., G.G., J.N. and Y.S. All authors have read and agreed to the published version of the manuscript.

## References

[1] Wilson, M. E.; Chen, L. H. Travellers Give Wings to Novel Coronavirus (2019-NCoV). Journal of Travel Medicine 2020, 27, taaa015. https://doi.org/10.1093/jtm/taaa015.

[2] Backer, J. A.; Klinkenberg, D.; Wallinga, J. Incubation Period of 2019 Novel Coronavirus (2019-NCoV) Infections among Travellers from Wuhan, China, 20-28 January 2020. Eurosurveillance 2020, 25. https://doi.org/10.2807/1560-7917.ES.2020.25.5.2000062.

[3] Linton, N.M.; Kobayashi, T.; Yang, Y.; Hayashi, K.; Akhmetzhanov, A.R.; Jung, S.-M.; Yuan, B.; Kinoshita, R.; Nishiura, H. Incubation Period and Other Epidemiological Characteristics of 2019 Novel Coronavirus Infections with Right Truncation: A Statistical Analysis of Publicly Available Case Data. J. Clin. Med. 2020, 9, 538. https://doi.org/10.3390/jcm9020538.

[4] Lauer, S. A.; Grantz, K. H.; Bi, Q.; Jones, F. K.; Zheng, Q.; Meredith, H. R.; Azman, A. S.; Reich, N. G.; Lessler, J. The Incubation Period of Coronavirus Disease 2019 (COVID-19) From Publicly Reported Confirmed Cases: Estimation and Application. Ann Intern Med 2020, 172, 577-582. https://doi.org/10.7326/M20-0504.

[5] Guan, W.; Ni, Z.; Hu, Y.; Liang, W.; Ou, C.; He, J.; Liu, L.; Shan, H.; Lei, C.; Hui, D. S. C.; et al. Clinical Characteristics of Coronavirus Disease 2019 in China. N Engl J Med 2020, 382, 1708-1720. https://doi.org/10.1056/NEJMoa2002032.

[6] Kong, T.K. Longer incubation period of coronavirus disease 2019 (COVID-19) in older adults. Aging Medicine 2020.

[7] Jiang, A. B.; Lieu, R.; Quenby, S. Significantly Longer Covid-19 Incubation Times for the Elderly, from a Case Study of 136 Patients throughout China. *medRxiv* 2020.

[8] Yang, L.; Dai, J.; Zhao, J.; Wang, Y.; Deng, P.; Wang, J. Estimation of Incubation Period and Serial Interval of COVID-19: Analysis of 178 Cases and 131 Transmission Chains in Hubei Province, China. Epidemiology and Infection 2020, 148, e117.

[9] DXY: COVID-19 Epidemic situation in real time. Available online: https://docs.google.com/spreadsheets/d/e/2PACX-1vQU0SIALScXx8VXDX7yKNKWWPKE1YjFlWc6VTEVSN45CklWWf-uWmprQIyLtoPDA18tX9cFDr-aQ9S6/pubhtml (accessed on 20 March 2020).

[10] Xu, B.; Gutierrez, B.; Mekaru, S.; Sewalk, K.; Goodwin, L.; Loskill, A.; Cohn, E. L.; Hswen, Y.; Hill, S. C.; Cobo, M. M.; et al. Epidemiological Data from the COVID-19 Outbreak, Real-Time Case Information. Sci Data 2020, 7, 106. https://doi.org/10.1038/s41597-020-0448-0.

[11] Dabrowska, D. M.; Doksum, K. A. Estimation and Testing in a Two-Sample Generalized Odds-Rate Model. Journal of the American Statistical Association 1988, 83, 744-749. https://doi.org/10.1080/01621459.1988.10478657.

[12] R Core Team (2019). R: A language and environment for statistical computing. R Foundation for Statistical Computing, Vienna, Austria. URL https://www.R-project.org/.

[13] Cowling, B. J.; Muller, M. P.; Wong, I. O. L.; Ho, L.-M.; Louie, M.; McGeer, A.; Leung, G. M. Alternative Methods of Estimating an Incubation Distribution: Examples from Severe Acute Respiratory Syndrome. Epidemiology 2007, 18, 253-259.

[14] World Health Organization (WHO). Considerations for quarantine of individuals in the context of containment for coronavirus disease (COVID-19). Available online: https://www.who.int/publications-detail/considerations-forquarantine-of-individuals-in-the-context-of-containment-forcoronavirus-disease-(covid-19) (accessed on 12 April 2020)

[15] The White House. Press Briefing by Members of the President’s Coronavirus Task Force. Available online: https://www.whitehouse.gov/briefings-statements/press-briefing-members-presidents-coronavirus-task-force (accessed on 1 February 2020).

[16] Moons K. G.; Altman D. G.; Reitsma J. B., et al. Transparent Reporting of a multivariable prediction model for Individual Prognosis or Diagnosis (TRIPOD): explanation and elaboration. Annals of internal medicine 2015, 162(1), Wl-73. 2015.

[17] Royston P.; Altman D. G.; Sauerbrei W. Dichotomizing continuous predictors in multiple regression: a bad idea. Stat. Med. 2006, 25, 127-141. 2015.

[18] Chowdhury, M.A.; Hossain, N.; Kashem, M.A.; Shahid, M.A.; Alam, A. Immune response in COVID-19: A review. Journal of Infection and Public Health 2020.

[19] van Dorp, L.; Acman, M.; Richard, D.; Shaw, L. P.; Ford, C. E.; Ormond, L.; Owen, C. J.; Pang, J.; Tan, C. C. S.; Boshier, F. A. T.; Ortiz, A. T.; Balloux, F. Emergence of Genomic Diversity and Recurrent Mutations in SARS-CoV-2. Infection. Genetics and Evolution 2020, 83, 104351.

